# Matching Adjusted Indirect Comparisons (MAICs) and Systematic Review: Efficacy and Safety of Experimental Chimeric Antigen Receptor (CAR) T-cells versus Axicabtagene ciloleucel (Yescarta) for the Treatment of Relapsed/Refractory Large B-Cell Lymphoma (LBCL)

**DOI:** 10.1101/2021.10.24.21265450

**Authors:** Bayarmagnai Weinstein, Bogdan Muresan, Sara Solano, Antonio Vaz de Macedo, YoonJung Lee, Yu-Chen Su, Yeseul Ahn, Gabriela Henriquez, Cristina Camargo, Gwang-Jin Kim, David O. Carpenter

**Affiliations:** Principles and Practice of Clinical Research (PPCR) Program, ECPE, Harvard T.H. Chan School of Public Health, Boston, MA 02115, USA; Department of Environmental Health Sciences, School of Public Health, University at Albany, Rensselaer, NY 12144, USA; Health Sciences Unit, University of Groningen, University Medical Center Groningen, Groningen, The Netherlands; Hematology Clinic, Hospital da Polícia Militar, Belo Horizonte, MG, CEP 30110-013, Brazil; Department of Pharmaceutical Sciences, School of Pharmacy, Texas Tech University Health Sciences Center, Amarillo, TX 79106, USA; Inari Medical, Biostatistics and Programming Dept, Irvine, CA 92618, USA; Iberoamerican University, Santo Domingo 10203, Dominican Republic; School of Medicine, Univesida de São Paulo, Brazil CEP 01246903; Institute of Experimental and Clinical Pharmacology and Toxicology, Faculty of Medicine, University of Freiburg, 79104 Freiburg, Germany; Institute for Health and the Environment, University at Albany, Rensselaer, NY 12144, USA

## Abstract

Despite favorable results of CAR T-cell therapy for relapsed/refractory large B-cell lymphoma (R/R LBCL), several challenges remain, including incomplete response, immune-mediated toxicity, and antigen-loss relapse. We delineated the relative clinical benefit of the novel approaches compared to the currently approved CAR T-cell therapies. In the absence of head-to-head comparisons and randomized controlled trials, we performed Matching Adjusted Indirect Comparisons to quantify the relative efficacy and safety of experimental CARs against Axicabtagene ciloleucel (Yescarta), the first FDA-approved CAR. A total of 182 R/R LBCL patients from 15 clinical trials with individual patient data (IPD) were pooled into eight populations by their CAR T-cell constructs and +/- ASCT status. The study endpoints were Progression-Free Survival (PFS), grade ≥ 3 cytokine release syndrome (CRS), and grade ≥ 3 neurotoxicity (NT). Tandem CD19.CD20.4-1BBζ CARs indicated favorable efficacy and safety, whereas the co-infusion of CD19 & CD20 with 4-1BBζ showed no clinical benefit compared to Yescarta. Third generation CD19. CD28. 4-1BBζ, and sequential administration of autologous stem cell transplantation (ASCT) and CD19. CARs presented statistically insignificant yet improved PFS and safety except for ASCT combined intervention which had suggestively higher NT risk than Yescarta. CARs with modified co-stimulatory domains to reduce toxicity (Hu19. CD8.28Zζ and CD19. BBz.86ζ) presented remarkable safety with no severe adverse events; however, both presented worse PFS than Yescarta. Third-generation CARs demonstrated statistically significantly lower NT than Yescarta. CD20. 4-1BBζ data suggested targeting CD20 antigen alone lacks clinical or safety benefit compared to Yescarta. Further comparisons with other FDA-approved CARs are needed.

**NOVELTY AND IMPACT:** Although currently approved CAR T-cells demonstrated unprecedently high response in relapsed / refractory LBCL in the salvage setting, lack of outcome durability and toxicity remain. We delineated the relative clinical benefit of the innovative experimental CAR T-cell approaches to Yescarta for insights into the ongoing efforts to address these inadequacies. Tandem CAR T-cells may provide higher efficacy and safer profile than Yescarta. Toxicity attenuated CAR T-cells present remarkable safety but no Progression-Free Survival (PFS) benefit.

## INTRODUCTION

Large B-cell lymphomas (LBCL) comprise diverse types of B-cell Non-Hodgkin Lymphoma (NHL), of which diffuse large B-cell lymphoma (DLBCL) is the most common histologic subtype, accounting for approximately a quarter of NHL cases worldwide (1). Survival rates have greatly improved over the past decades, particularly in the immunochemotherapy era, with a 5-year relative survival rate reported between 55.4% and 62.0% in developed countries (2). However, despite the advances achieved with rituximab-based regimens, up to 50% of patients with advanced-stage de novo DLBCL, for instance, will eventually relapse, even after achieving a complete response (CR) (3). If progression occurs during the initial treatment phase or soon after a brief CR, only 30% to 40% will respond to salvage chemotherapy and will be able to undergo consolidation with autologous stem cell transplantation (ASCT) (4). Even so, among these patients, roughly half will ultimately relapse after transplantation (5). The prognosis, in such cases, is poor, especially for those who have high-risk factors or relapse within 12 months post-ASCT (4,5). Thus, effective treatment for R/R LBCL remains a highly unmet need.

To date, only three CAR T-cell products (axicabtagene ciloleucel (Axi-cel, Yescarta), tisagenlecleucel (Tisa-cel, Kymriah), lisocabtagene maraleucel (Liso-cel, Breyanzi) are approved by the FDA for R/R LBCL (6–8). Despite the unprecedently high efficacy of these CAR T-cell therapies compared to historical outcomes for patients with R/R LBCL, current challenges, such as incomplete response, immune-mediated toxicity, and post-treatment relapse, remain. For example, in the ZUMA-1 trial for R/R LBCL, only 39% of patients maintained a CR to the therapy at the median of 27-month follow-up despite the initially high (82%) objective response rate (ORR) achieved (7,9). In an attempt to optimize CAR T-cell characteristics to address these inadequacies, pre-clinical researches identified tumor antigen escape and CD19 antigen downregulation as potential causal factors for the suboptimal response and relapse observed after CAR T-cell therapy (10). Tumor antigen escape leads to low antigen density via transfer of target antigens from the tumor cells to the CAR T-cells. This process, known as trogocytosis, has been observed with CD19, CD22, mesothelin, and B-cell maturation antigen (BCMA) (11).

Existing evidence prior to this study encompasses diverse strategies focused on advancing CAR T-cell performance. Specific approaches already notable for both their feasibility and clinical and safety benefit include (i) Multi-antigen targeting CAR T-cells obtained through co-infusion or sequential administration of single-targeted CAR T-cells against different antigens. Alternatively, tandem and bicistronic constructs expressing two different CARs on a single or a separate chimeric protein(s), respectively (12); (ii) Third and advanced generation CAR T-cells using integrated co-stimulatory domains (13–15); (iii) Enhanced co-stimulatory domains intended at reducing toxicity and preserving potency (16,17); (iv) Combination therapy of CAR T-cells and immune checkpoint inhibitors (18); (v) Co-administration of ASCT and CAR T-cells (19–21); (vi) Alternative antigen targetings (other than CD19), such as CD20, CD22, CD27, ICOS, and OX40 (22).

### Summary of the evidence prior to this study

Dual targeting CAR T-cells: Preclinical studies demonstrated high anti-tumor potency with tandem CD19/CD20 CAR T-cells (23,24), sequential infusion of CD19 and CD79b CARs (25), co-infusion of CD19 and CD38 CAR T-cells (26), and CD19/CD37 constructs (27). The clinical benefit of tandem CD19/CD20 CARs (28–30), co-infusion of CD19 and CD20 CARs (31), and mixed infusions of CD22 and CD19 CAR T-cells (32,33) has been evaluated in small early phase clinical trials with demonstrated feasibility and varying levels of efficacy and safety.

Among the next-generation CAR T-cells, more mature data exist for the third-generation CAR T-cells incorporating both CD28ζ and 4-1BBζ co-stimulatory signaling domains. In mice models, third-generation CAR T-cells demonstrated improved T-cell persistence and stronger antitumor potency compared to second-generation constructs (34). In addition, the clinical benefits of third-generation CARs in LBCL patients were evaluated in early phase trials (13–15). However, whether the addition of 4-1BBζ co-stimulatory domains to a common CD28ζ domain enhances such clinical benefits compared to second-generation CAR T-cells in this population is still unclear.

Variations of CAR T-cells with modified co-stimulatory domains aimed at reducing treatment-related toxicity include (1) Hu19. CD8.28Z, containing a fully human single-chain variable fragment (scFv) and CD8α-based hinge and transmembrane domains (16); (2) CD19. BBz.86, with an 86-amino-acid fragment from human CD8α (17); and. Both CD19. BBz.86 and Hu19. CD8.28Z CAR T-cells demonstrated exceptional safety, yet attenuated efficacy, based on the CR rates of 29% and 39% observed, respectively, compared to the 54% CR rate noted among the LBCL patients receiving Axi-cel (Yescarta).

ASCT and CAR T-cell therapy: Multi-center randomized clinical trials are underway to determine the comparative efficacy and safety of CAR T-cell therapy alone vs. ASCT combined with systemic therapies for the treatment of R/R LBCL (refer to Discussion section for additional details). The study compared locally manufactured CD19. CD28ζ CAR T-cells in China to ASCT alone (NCT03196830) demonstrated superior efficacy and safety of the CAR T-cell product compared to ASCT in R/R NHL patients (35). Whether the sequential administration of ASCT and CAR T-cells hold higher clinical benefits than CAR T-cells alone remains to be elucidated. Of note, this has already been shown to be feasible and safe in three clinical trials (19–21).

CD20. 4-1BBζ CAR-T cells demonstrated high antitumor activity against LBCL in pre-clinical studies (36), and few clinical trials tested second-generation CD20 CAR T-cells in this disease (36–38). Clinical trials evaluating third-generation CD20 CARs are currently underway in China (NCT02710149), in the USA (NCT03277729) - evaluating MB-106, a fully human third-generation CD20.4-1BBζ.CD28 ζ CAR T-cell constructs - and in Germany (NCT03664635), with MB-CART20.1 CARs (39,40). Targeting CD20 was shown to be exceptionally more efficacious in follicular lymphoma, as demonstrated with the success of rituximab, an anti-CD20 monoclonal antibody, which led to the current rituximab-based first-line combination treatment for most NHL types (41). The fact that 30–40% of LBCL patients relapse after rituximab suggests that targeting CD20 alone is not enough (41). This has set the basis for comparative insights between CD20 and CD19-targeted CAR T-cells, thereby shedding light on the development of the dual targeting approaches mentioned above.

Even though such key pre-clinical and clinical data became available regarding the innovative approaches aiming to extending the durability of response beyond the achievements of the currently approved CAR T-cells, there is no comparative efficacy and safety data exist to shed a light on the relative advantages of the experimental CAR T-cell products versus the currently approved CAR T-cell therapies. Hence, we aimed to compare the efficacy and safety of the currently available experimental CAR T-cell products to Yescarta, the first FDA-approved CAR T-cell therepay, thereby harboring the longest follow-up data available to date. Also, to overcome limitations of all currently available CAR T-cell trials being single-arm trials, and individual patient-level data only available for experimental CAR T-cell products and not for ZUMA-1 trial, the comparator (42), we used unanchored matching-adjusted indirect comparison (MAIC) as a primary method. The MAIC techniques attenuate bias in comparing multiple treatments assessed in different studies by matching patient-level data from the clinical trials of one treatment to aggregate data by comparator trials. Aditionally, MAIC provides a more robust adjustment for cross-trial differences in patient characteristics than traditional meta-regressions due to its higher accuracy obtained from individual patient data than from aggregate data (43). We believe this systematic review-based quantitative comparison may provide guiding insights into the ongoing efforts to advance CAR T-cell therapy for the treatment of R/R LBCL.

## METHODS

### Data sources

For Yescarta, published aggregate data was used from the ZUMA-1 trial (disease- and baseline characteristics data) evaluating the efficacy of Yescarta for treatment of LBCL patients (7). For experimental CAR T-cell products for comparator arms, individual patient data (IPD) was available from the corresponding peer-reviewed publications that we identified through a Preferred Reporting Items for Systematic Reviews and Meta-Analyses (PRISMA) guideline-based systematic review. This study included the clinical trials for the patients who received CAR T-cell therapies for the treatment of RR-LBCL after two or more systemic therapies regardless of the type of CAR T-cells, geography, health care settings (inpatient and outpatient), and demographic characteristics (age, gender, race, or ethnicity). Clinical trials that provided concomitant therapies together with CAR T-cell products (except for bridging or lymphodepleting chemotherapy) were excluded. We searched the extensive scope of electronic databases including PubMed, Cochrane Central, Medline via Ovid, Embase via Ovid, Scopus Elsevier, Web of Science, and Education Resources Center (ERIC). Conference proceedings were identified from the websites of the American Society of Hematology, American Society of Clinical Oncology, and European Hematology Association. North American and international trials were ascertained through a search in ClinicalTrials.gov, International Standard Randomized Controlled Trial Number (ISRCTN registry), World Health Organization International Clinical Trials Registry Platform (ICTRP), Deutsches Register Klinischer Studien (DRKS), Chinese Clinical Trial Registry (ChiCTR), European Clinical Trials Register (www.clinicaltrialsregister.eu), Latin American and Caribbean Health Science Information Database (LILACS), and Australian and New Zealand Clinical Trials Registry. To identify potential publication bias, we searched for unpublished trials and grey literature using Google, Grey Literature Report (greylit.org), and OpenGrey (opengrey.eu).

To reduce a potential confounding to the study results due difference in the study designs that MAIC method is not designed to address, we conducted a feasibility assessment and Risk of Bias Assessment, NIH Quality Assessment Tool for Case Series Studies (44) by comparing the eligible studies in terms of their PICOS (population, intervention, comparator, outcomes, and study design) and the NIH recommended criteria. Consequently, we excluded four studies with significantly different PICOS from the rest of the studies (Figures S1). Details regarding the literature search terms and queries, study selection, data extraction, and the risk of bias assessment can be found in the published protocol for the present study (45).

We created eight independent interventions of distinct types of CAR T-cell products where five of them were pooled population from multiple trials evaluating the similar CAR T-cell constructs as shown in Table 1 and listed as follows:

**Table 1.**
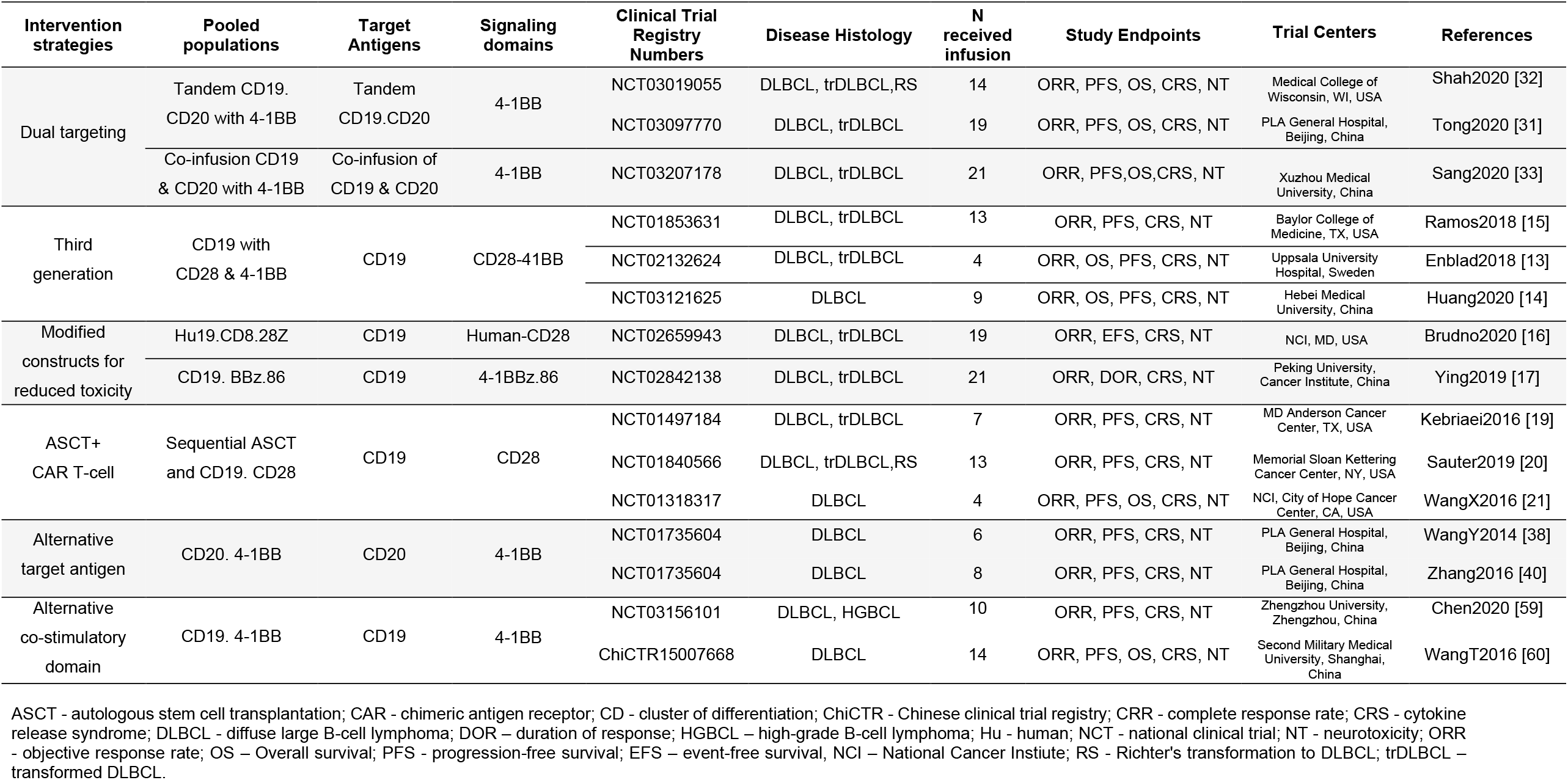
Summary of Clinical Trials, Pooled Populations by CAR T-cell structure, and Study Endpoints.

1. Dual targeting using tandem CD19. CD20 with 4-1BBζ, a pool of two trials
2. Dual targeting by co-infusion of CD19 & CD20 with 4-1BBζ
3. Third-generation CARs: CD19 with CD28ζ & 4-1BBζ, a pool of three trials
4. Modified co-stimulatory domain for reduced toxicity: Hu19.CD8.28Z
5. Modified co-stimulatory domain for reduced toxicity: CD19.BBz.86
6. Sequential administration of ASCT and CD19.CD28ζ, a pool of three trials
7. Alternative target-antigen: CD20. 4-1BBζ CARs, a pool of two trials
8. Alternative co-stimulatory domain: CD19. 4-1BBζ CARs in Chinese patients, a pool of two trials

Pooling patients when possible have augmented statistical power and also enabled us to test the hypothesis by CAR T-cell types. We excluded the trials with less than 10 patients (Figure S1). The trials for CAR T-cells that eventually evolved into Yescarta, and any early phase trials of the CAR T-cells developed into Kymriah and Breyanzi, currently approved products, were also excluded from this study.

Reconstructed patient-level progression-free survival (PFS) data for ZUMA-1 trial: for the calculation of the hazard ratio (HR) and its 95% confidence interval (CI) associated with the PFS of each pooled CAR T-cell population versus Yescarta, we reconstructed individual patient PFS data from the ZUMA-1 trial through a validated algorithm developed by Guyot and colleagues (2012) (46). This was achieved by obtaining the number of patients at risk and the total number of events along with the geometric coordinates of the published PFS Kaplan-Meier (KM) curve associated with Yescarta over a 24-month follow-up time using Origin digitizing software.

### Outcomes assessed

We selected PFS for efficacy and grade ≥ 3 cytokine release syndrome (CRS) and neurotoxicity (NT) for safety outcomes, given that the purpose of the study was to determine the relative benefit of experimental CAR T-cell products compared to Yescarta in terms of response durability and severe toxicity. Table 1 presents the study endpoints reported by the eligible studies. Overall survival (OS) was not used since multiple studies had not reached the median follow-up time at the time of this analysis. Likewise, we did not focus on the objective response rate or the initial response types, as these measures do not directly reflect the durability of response over time, and there is notable cross-trial variation in the timing for the objective response measurements.

### Statistical methods

Given the existing evidence limited to single-arm trials, we conducted unanchored MAICs to adjust for cross-trial heterogeneity in baseline characteristics. Matching covariates were selected following the NICE Guidelines given a) mutually reported disease- and patient baseline characteristics, where the feasibility assessment revealed cross-trial heterogeneity, b) whose clinical meaningfulness was confirmed by clinicians/experts (Table 2).

**Table 2.**
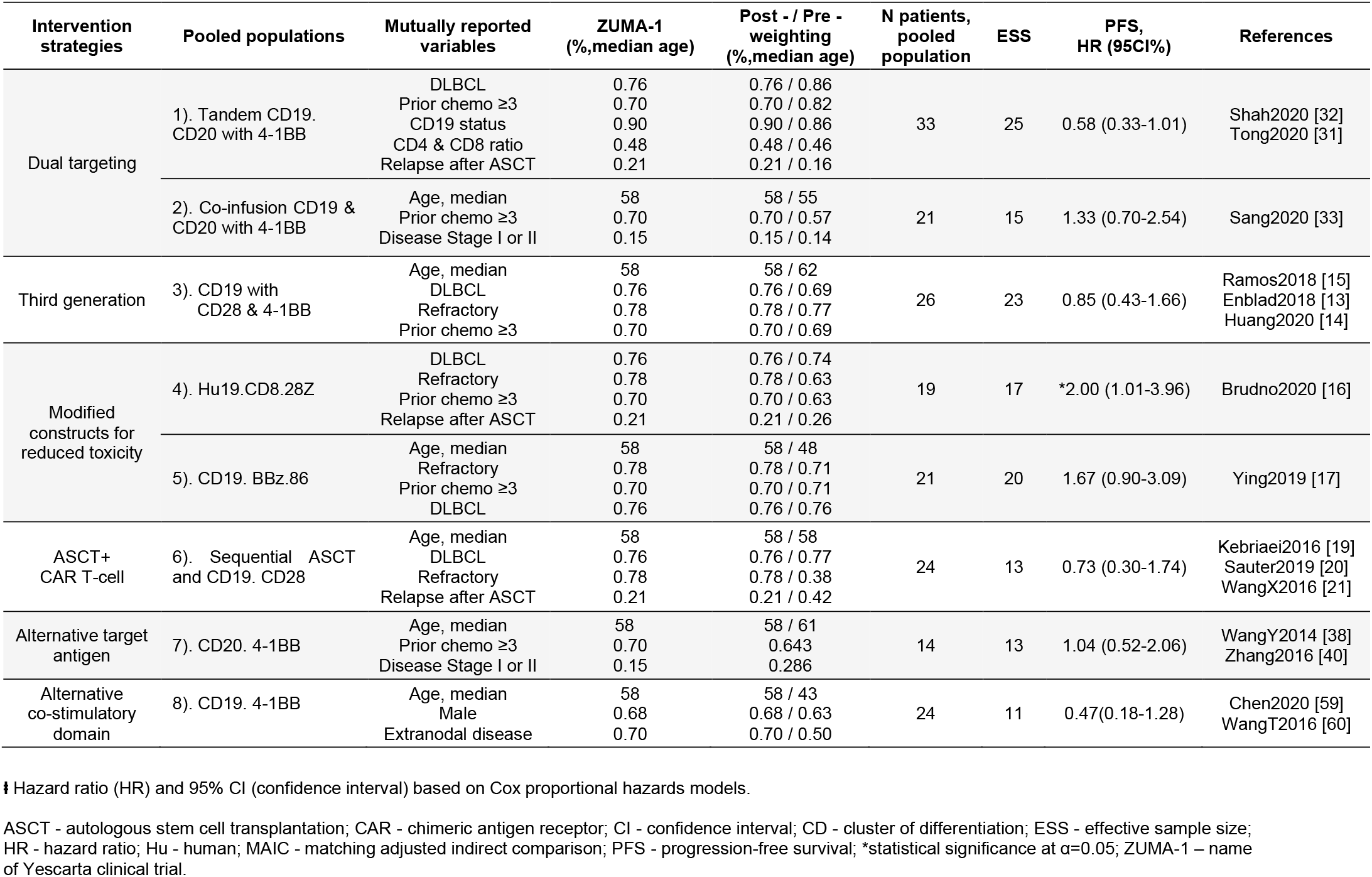
Key Baseline Characteristics and MAICs of Experimental CAR T-cells versus Yescarta regarding Progression-Free Survival ⱡ.

| Intervention strategies | Pooled populations | Mutually reported variables | ZUMA-1<br>(%,median age) | Post - / Pre -<br>weighting<br>(%,median age) | N patients,<br>pooled<br>population | ESS | PFS,<br>HR (95CI%) | References |
| --- | --- | --- | --- | --- | --- | --- | --- | --- |
| Dual targeting | 1). Tandem CD19.<br>CD20 with 4-1BB | DLBCL<br>Prior chemo ≥3<br>CD19 status<br>CD4 & CD8 ratio<br>Relapse after ASCT | 0.76<br>0.70<br>0.90<br>0.48<br>0.21 | 0.76 / 0.86<br>0.70 / 0.82<br>0.90 / 0.86<br>0.48 / 0.46<br>0.21 / 0.16 | 33 | 25 | 0.58 (0.33-1.01) | Shah2020 [32]<br>Tong2020 [31] |
|  | 2). Co-infusion CD19 &<br>CD20 with 4-1BB | Age, median<br>Prior chemo ≥3<br>Disease Stage I or II | 58<br>0.70<br>0.15 | 58 / 55<br>0.70 / 0.57<br>0.15 / 0.14 | 21 | 15 | 1.33 (0.70-2.54) | Sang2020 [33] |
| Third generation | 3). CD19 with<br>CD28 & 4-1BB | Age, median<br>DLBCL<br>Refractory<br>Prior chemo ≥3 | 58<br>0.76<br>0.78<br>0.70 | 58 / 62<br>0.76 / 0.69<br>0.78 / 0.77<br>0.70 / 0.69 | 26 | 23 | 0.85 (0.43-1.66) | Ramos2018 [15]<br>Enblad2018 [13]<br>Huang2020 [14] |
| Modified<br>constructs for<br>reduced toxicity | 4). Hu19.CD8.28Z | DLBCL<br>Refractory<br>Prior chemo ≥3<br>Relapse after ASCT | 0.76<br>0.78<br>0.70<br>0.21 | 0.76 / 0.74<br>0.78 / 0.63<br>0.70 / 0.63<br>0.21 / 0.26 | 19 | 17 | *2.00 (1.01-3.96) | Brudno2020 [16] |
|  | 5). CD19. BBz.86 | Age, median<br>Refractory<br>Prior chemo ≥3<br>DLBCL | 58<br>0.78<br>0.70<br>0.76 | 58 / 48<br>0.78 / 0.71<br>0.70 / 0.71<br>0.76 / 0.76 | 21 | 20 | 1.67 (0.90-3.09) | Ying2019 [17] |
| ASCT+<br>CAR T-cell | 6). Sequential ASCT<br>and CD19. CD28 | Age, median<br>DLBCL<br>Refractory<br>Relapse after ASCT | 58<br>0.76<br>0.78<br>0.21 | 58 / 58<br>0.76 / 0.77<br>0.78 / 0.38<br>0.21 / 0.42 | 24 | 13 | 0.73 (0.30-1.74) | Kebriaei2016 [19]<br>Sauter2019 [20]<br>WangX2016 [21] |
| Alternative target<br>antigen | 7). CD20. 4-1BB | Age, median<br>Prior chemo ≥3<br>Disease Stage I or II | 58<br>0.70<br>0.15 | 58 / 61<br>0.643<br>0.286 | 14 | 13 | 1.04 (0.52-2.06) | WangY2014 [38]<br>Zhang2016 [40] |
| Alternative<br>co-stimulatory<br>domain | 8). CD19. 4-1BB | Age, median<br>Male<br>Extranodal disease | 58<br>0.68<br>0.70 | 58 / 43<br>0.68 / 0.63<br>0.70 / 0.50 | 24 | 11 | 0.47(0.18-1.28) | Chen2020 [59]<br>WangT2016 [60] |

‡ Hazard ratio (HR) and 95% CI (confidence interval) based on Cox proportional hazards models.
ASCT - autologous stem cell transplantation; CAR - chimeric antigen receptor; CI - confidence interval; CD - cluster of differentiation; ESS - effective sample size; HR - hazard ratio; Hu - human; MAIC - matching adjusted indirect comparison; PFS - progression-free survival; \*statistical significance at $\alpha=0.05$ ; ZUMA-1 – name of Yescarta clinical trial.

In MAIC, patients in experimental CAR T-cell trials with IPD were re-weighted to match the mean baseline characteristics in ZUMA-1 with only aggregate data. The weights were estimated by the method of moments, applied to the IPD, so the summary statistics of the baseline characteristics of the IPD becomes similar to those of the aggregate data (43). Based on the calculated weights, individual patient-level PFS and percentage of grade ≥3 CRS and NT were re-weighted for further survival and logistic regression analyses. For the comparator arm, reconstructed individual patient level PFS for Yescarta (see Materials section) was used. Given these data, Cox proportional hazards (PH) model estimated the HR and its 95% CI for PFS associated with each pair of eight pooled CAR T-cell populations versus Yescarta. Corresponding weighted KM curves were created assuming exact ties according to the Kalbfleisch-Prentice method. Finally, logistic regression models were used to compute the odds ratio (OR) and its 95% CI based on the re-weighted data for both safety outcomes: grade ≥ 3 CRS and NT.

A recent MAIC study of Yescarta vs. Kymriah identified the LBCL-specific key prognostic covariates and demonstrated refractory status and number of prior therapies as the most influential variables on the CAR T-cell treatment outcomes (47). We identified the mutually reported key baseline covariates and used categorizations as follows: age (<58 years), disease stage (<3), histology (diffuse LBCL/other types), refractory status, number of prior lines of therapy (>4), and extranodal disease status. The pack of mutually reported covariates varied for each pair of distinct pool CAR T-cell population and Yescarta, as this was dictated by the size of the IPD pooled population and the mutual availability of the data in both the IPD and ZUMA-1 trials, as shown in Table 2. The degree of overlap between pairwise comparisons reflects in ESS (Table 2). All analyses were performed using R version 4.1.0 (2021). The survival package, along with the necessary supporting functions, was used to estimate alternative survival functions by trial.

## RESULTS

Individual Patient Data (IPD): Through systematic review, we identified 15 clinical trials for experimental CAR T-cell products (Table 1) with individual patient data (IPD), as presented in the PRISMA flow diagram in Figure S1, Supplementary Materials. Table 2 presents the effective sample size (ESS) and the weighted versus unweighted values of matching baseline covariates across each pooled CAR T-cell population versus ZUMA-1.

### Dual targeting CARs versus Yescarta

#### Tandem CD19. CD20 with 4-1BB

Our MAIC weighted analysis showed that the tandem CD19.CD20.4-1BBζ CAR T-cells presented suggestive evidence of increased PFS (HR = 0.58; 95% CI, 0.33-1.01), reduced grade ≥ 3 CRS (OR=0.70; 95% CI, 0.18-2.76) and statistically significantly lower odds of grade ≥ 3 NT (OR=0.14; 95% CI, 0.02-0.78) compared to Yescarta (Table 2 & 3).

**Table 3.** MAICs of Experimental CAR T-cells versus Yescarta Regarding Grade ≥3 CRS and NT.

| Intervention strategy | Pooled populations | N received infusion | Sum of weights | Number of CRS, grade ≥ 3 | OR (95%CI) CRS, grade ≥ 3 | Number of NT, grade ≥ 3 | OR (95%CI) NT, grade ≥ 3 | References |
| --- | --- | --- | --- | --- | --- | --- | --- | --- |
| Dual targeting | 1). Tandem CD19. CD20. 4-1BB | 33 | 28 | 3 | 0.70 (0.18-2.76) | 2 | *0.14 (0.02-0.78) | Shah2020 [32]<br>Tong2020 [31] |
|  | 2). Co-infusion CD19 & CD20 with 4-1BB | 21 | NA | NA | NA | NA | NA | Sang2020 [33] |
| Third generation | 3). CD19 with CD28 & 4-1BB | 26 | 26 | 1 | 0.20 (0.02-2.12) | 1 | *0.20 (0.04-0.94) | Ramos2018 [15]<br>Enblad2018 [13]<br>Huang2020 [14] |
| Modified constructs for reduced toxicity | 4). Hu19. CD8.28Z | 14 | 20 | 0 | 0 | 0 | 0 | Brudno2020 [16] |
|  | 5). CD19. BBz.86 | 21 | 20 | 0 | 0 | 0 | 0 | Ying2019 [17] |
| ASCT+ CAR T-cell | 6). Sequential ASCT and CD19. CD28 | 24 | 16 | 3 | 0.25 (0.02-3.68) | 7 | 1.78 (0.60-5.28) | Kebriaei2016 [19]<br>Sauter2019 [20]<br>WangX2016 [21] |
| Alternative target antigen | 7). CD20. 4-1BB | 14 | 13 | 2 | 1.04 (0.20-5.38) | NA | NA | WangY2014 [38]<br>Zhang2016 [40] |
| Alternative co-stimulatory domain | 8). CD19. 4-1BB | 24 | 12 | 4 | 0.95 (0.15-5.94) | NA | NA | Chen2020 [59]<br>WangT2016 [60] |

ASCT - autologous stem cell transplantation; CAR - chimeric antigen receptor; CI - confidence interval; CD - cluster of differentiation; CRS - cytokine release syndrome; OR - odds ratio; Hu - human; MAIC - matching adjusted indirect comparison; \*statistical significance at α=0.05; NT - neurotoxicity; ZUMA-1 – name of Yescarta clinical trial.

#### Co-infusion CD19 & CD20 with 4-1BB

In contrast, co-infusion of CD19 and CD20 CAR-T cells had statistically insignificant but worse PFS (HR=1.33, 95% CI: 0.70-2.54) than Yescarta. IPD for safety outcomes was not available from this study. However, naïve direct comparison shows this co-infusion approach presented higher grade ≥ 3 CRS (28.5% vs. 13% in ZUMA-1) and lower neurotoxicity (9.5% vs. 28% in ZUMA-1) than Yescarta.

#### Third-generation CARs versus Yescarta

We found suggestive evidence of improved PFS with HR=0.85 (95% CI: 0.43-1.66) and safety in terms of grade ≥3 CRS with OR=0.20 (95% CI: 0.02-2.12) and NT with OR=0.20 (95% CI: 0.04-0.94) associated with third-generation CAR T-cells versus Yescarta.

#### Modified co-stimulatory domains for reduced toxicityversus Yecarta

Both Hu19.CD8.28Z and CD19. BBz.86 CAR T-cells presented excellent safety profiles with the absence of grade ≥3 CRS and NT. However, yet both of these CAR T-cells presented reduced benefit in PFS: Hu19.CD8.28Z with HR=2.00; 95% CI, 1.01-3.96 and CD19. BBz.86 with HR=1.67; 95% CI, 0.90-3.09 compared to Yescarta, though without statistical significance in CD19. BBz.86.

#### Sequential administration of ASCT and CD19. CD28ζ versus Yescarta

Our findings demonstrated suggestive favorable PFS (HR=0.73; 95% CI, 0.30-1.74) and reduced grade ≥ 3 CRS (OR=0.25; 95% CI, 0.02-3.68) but increased NT (OR=1.78; 95% CI, 0.60-5.28) for the sequential administration of CD19. CD28ζ CAR T-cells within 2 to 6 days after ASCT compared to Yescarta. None of these findings reached statistical significance.

#### Alternative target antigen: CD20. 4-1BBζ versus Yescarta

We did not find any notable clinical benefit or harm in this pooled population treated with CD20. 4-1BBζ versus Yescarta in terms of PFS (HR= 1.04; 95% CI, 0.52-2.06) and CRS (OR=1.04; 0.20-5.38). We didn’t examine the neurotoxicity as IPD was not available from this trial.

#### Alternative co-stimulary domain: CD19. 4-1BBζ in Chinese patients versus Yescarta

The MAIC of CD19. 4-1BBζ CARs based on the pooled population of two small trials conducted in China, Shanghai (48,49) versus Yescarta showed no significant difference but suggestively better PFS than that of Yescarta, with HR=0.47 (95% CI, 0.18-1.28) and slightly reduced grade ≥ 3 CRS (OR=0.95; 95% CI, 0.15-5.94) than Yescarta. These two trials did not provide neurotoxicity data.

## DISCUSSION

### Dual targeting strategies versus Yescarta

The improved PFS associated with tandem CAR T-cells whereas reduced PFS associated with co-infusion of CD19 and CD20 CAR T-cells versus Yescarta corroborates pre-clinical studies that demonstrated the higher efficacy and safety of tandem CAR T-cells than that of co-infusions (10). The reduced survival benefit and increased CRS associated with the co-infusion of different CAR T-cell targets may be associated with (1) additive toxicity from stronger cytokine storm through the amplified number of targetable antigens; (2) competitive targeting limit the expansion of other CAR T-cells; (3) compromised engraftment due to the interference of multiple antigens (12,50,51). Literature review on the multi-antigen targeting strategies show that there is more data for CD19 and CD22 covering various administration approaches (sequential and co-infusion) and different constructs (tandem and bicistronic). A study showed that sequential administration of CD19 and CD22 CAR T-cells in 12 DLBCL patients (32) and co-infusion in 36 NHL patients (33) resulted in objective response rates of 77% and 83% and grade ≥3 CRS rates of 14% and 21%, respectively. Tandem CD19 and CD22 CAR T-cells appeared to be feasible and potentially efficacious in R/R B-cell acute lymphoblastic leukemia (B-ALL) (52). Although these preliminary results are somewhat comparable to the 82% ORR and 13% grade ≥3 CRS observed in ZUMA-1, longer follow-up data are required to assess whether sequential and mixed infusion approaches reduce post-CAR T-cell therapy relapse. Bicistronic CD19. CD22 trials in pediatric and adult R/R B-ALL (53,54) demonstrated unprecedently high CR rates (100%) and a notable safety profile with a single occurrence of grade ≥3 CAR T-cell related encephalopathy syndrome (CRES) in the pediatric trial. As for B-cell lymphoma, bicistronic CD19.CD22 trials led by Shah and colleagues (NCT03448393), Miklos and colleagues (NCT03233854), and Pulsipher and colleagues (NCT03330691) are currently in progress.

### Third-generation CARs versus Yescarta

Although the result associated with the third-generation CAR T-cells versus 15escarta was not statistically significant, the slight protective effects observed may be due to the multifunctional cytokine secretion and improved persistence of T-cells from the concurrent expression of CD28ζ and 4-1BBζ co-stimulatory domains versus that of the CD28ζ co-domain alone (55). In addition, in-vivo studies demonstrated that adding 4-1BBζ to the second-generation construct protects CD28ζ tumor-specific cells from activation-induced cell death while supporting central memory cells and mitochondrial functions (56). In alignment with this existing evidence, three contributing IPD trials of the pooled population in our study reported high expansion and improved persistence of T-cells in common. Moreover, all three trials of third-generation CAR T-cells analyzed in this study have highlighted that the patients with less tumor burden and who were prior responders to chemotherapy had higher tumor clearance benefits than patients with more tumor burden and non-responders to chemotherapy (13–15).

### Modified co-stimulatory domains for reduced toxicity

Hu19.CD8.28Z and CD19. BBz.86 CAR T-cells were designed to exert minimal toxicity while preserving antitumor potency to Yescarta. Reduced cytokine-mediated toxicity is often accomplished by attenuating CAR signal strength and enhancing T-cell persistence (16,17,57). Consequently, this enables tumor immune escape and hampers the antitumor potency of CAR T-cells, particularly for low antigen density tumors (58). This may explain our findings of lower toxicity and reduced PFS benefit of two anti-CD19 CAR T-cells (Hu19.CD8.28Z and CD19. BBz.86). Multiple pre-clinical studies are underway toward determining the CAR structure that might achieve minimum toxicity and maximum efficacy for low antigen density tumors. A recent in-vivo leukemia model demonstrated the high potency of a new CD19. CD28ζH/T-4-1BBζ construct, despite the low antigen density of the leukemic cells, while accomplishing a similar efficacy to Yescarta (58). Further studies are required to assess how these pre-clinical findings translate into clinical benefits for lymphoma patients.

#### Sequential administration of ASCT + CD19. CD28ζ versus Yescarta

The authors of the clinical trials that evaluated the safety and efficacy of the sequential administration of ASCT and CD19. CD28ζ CAR T-cell therapy in LBCL patients (19–21) hypothesized that this combination will reduce cytokine production while exerting a high antitumor potency by increasing the expansion and persistence of CAR T-cells. Providing ASCT prior to CAR T-cell administration is believed to reduce tumor burden, diminish immuno-suppressive microenvironment, and boost lymphodepletion, thereby reducing the number of regulatory T-cells and myeloid cells. Nevertheless, our findings on ASCT + CD19. CD28ζ versus Yescarta were inconclusive as none of the findings reached statistical significance. The direction of the results remained unchanged in separate analyses for the U.S. and Chinese trials. Nevertheless, the feasibility and clinical benefit of the concurrent administration of ASCT with CAR T-cell therapy may not be justifiable since this combined regimen would not be available for about half of the patients who are transplant-ineligible due to chemo-refractory disease and half of those who received ASCT yet still at risk for disease relapse post-autografting (59). Of note, combining ASCT with CAR T-cell therapy may not be necessary if CAR T-cell therapy alone is superior to ASCT, as previously shown (35). Intensive efforts are underway to understand whether CAR T-cell therapy is efficacious and safe to replace ASCT in earlier lines of treatment of LBCL. A few randomized multi-center clinical trials are underway comparing the FDA-approved CAR T-cells - Yescarta in the ZUMA-7 (NCT03391466), Kymriah in the BELINDA (NCT03570892), and Breyanzi in the TRANSFORM (NCT03575351) trials versus the standard of care comprised of systemic therapies followed by ASCT.

#### Alternative target-antigen: CD20. 4-1BBζ versus Yescarta

CD19 has been a primary target in CAR T-cell therapy for LBCL due to its pan B-cell expression and increased expression in B-cell leukemias and lymphomas (60). In contrast, CD20 and CD22 have limited expression in mature B cells. Nevertheless, targeting both CD19 and CD20 has an additive effect, given CD20 antigen’s higher average density of surface molecules per tumor cell, combined with CD19’s pan B-cell lineage cell expression, with extended-expression in certain CD20-negative tumor subsets (61). Since all patients in this trial were treated with rituximab before CD20.4-1BBζ CAR T-cell administration, the question of whether CD20. CAR T-cells would be more efficacious among rituximab-näive patients remain to be clarified.

#### Alternative co-stimulatory domain: CD19. 4-1BBζ in Chinese patients versus Yescarta

This CAR T-cell product has identical construct with Tisagenlecleucel (Kymriah) except the trial was conducted in different study population in China. Despite the statistical non-significance of the findings for the CD19. 4-1BBζ in Chinese patients versus Yescarta comparison in this study, the slight improvement in safety was associated with the CD19. 4-1BBζ CAR T-cells versus Yescarta in these studies is consistent with the recent MAIC of Kymriah and Breyanzi to Yescarta (47). 4-1BBζ is one of the well-established co-stimulatory domains incorporated into the CD19 CARs in Kymriah and Breyanzi, while Yescarta contains a CD28ζ co-domain. The impact of the CD28ζ versus 4-1BBζ co-stimulatory domains on CAR T-cell behavior has been studied in in-vivo studies and multiple clinical studies in B-ALL. CD19. CD28ζ CAR T-cells show a faster and higher peak expansion, yet reduced T-cell persistence compared to 4-1BBζ-containing CARs (62). Nonetheless, it is not fully clear whether CAR T-cell persistence is a strong determinant of response durability in LBCL as it is for B-ALL.

#### Comparative effectiveness studies on currently approved CAR T-cell therapies

A recently published MAIC of Yescarta versus Kymriah demonstrated superior efficacy of Yescarta, with higher CR rates (RR=1.62, 95% CI: 1.16-2.27) and improved OS (HR=0.51, 95% CI: 0.31-0.83), yet increased toxicity, with grade 1-2 CRS with OR = 6.20 (95% CI: 2.76-13.93) and grade ≥ 3 NT with OR=2.20 (95% CI: 0.98-3.60) in R/R LBCL (47). Another recently published MAIC of Yescarta versus Breyanzi demonstrated similar efficacy, slightly favoring Yescarta (PFS with HR=1.30; 95%, 0.96-1.77). However, Breyanzi presented a significantly safer profile than Yescarta (grade ≥3 CRS and NT with OR= 0.16; 95% CI, 0.06-0.47, and OR=0.31, 95% CI, 0.18-0.54, respectively (63). Among the currently approved CAR T-cells, based on the existing MAICs, Yescarta appears to present higher efficacy than Kymriah and comparable efficacy to Breyanzi. In contrast, the latter two incorporating 4-1BBζ co-domains demonstrate a safer profile regarding CRS and NT than Yescarta. The lower toxicity and similar efficacy observed with Breyanzi versus Yescarta relates to its ability to induce a low variability in cytokine production (IL-2, IFN-γ, TNF-α, etc.). This was accomplished through controlled manufacturing to maintain the ratio of CD4+ and CD8+ to 1:1 under optimized culture conditions. In-vivo studies are underway to clarify the exact underlying mechanisms in this regard (64).

### Strengths, Limitations, and Future Study

To our knowledge, this study is the first to report the indirect comparison of experimental CAR T-cells to Yescarta, an FDA-approved CAR T-cell product. This study incorporated a systematic literature review with MAICs, the only statistical tool to address studies in the absence of direct head-to-head comparisons and the presence of single-arm trials only. Consequently, we were able to systematically retrieve comprehensive data from an IPD bank of experimental CAR T-cells.

The results of this study need to be evaluated in light of a number of important limitations. Even though we attempted to account for cross-trial heterogeneity using MAICs, it is important to acknowledge the residual case-mix and beyond case-mix heterogeneity (65). This means that the study results are subject to residual confounding since MAICs can only correct for heterogeneity in mutualy reported disease- and patient baseline characetistics. Inevitable differences between the eligible clinical trials in terms of trial management strategies, study designs, protocols, presence or absence of conditioning regimes and/or bridging therapies, CAR T-cell engineering techniques, and manufacturing processes potentially introduces the bias in indirect comparison studies. For example, ZUMA-1 did not use bridging therapy, as opposed to some of the eligible IPD trials in this study used bridging therapy. It is unclear as to how this may have affected our results since the role of bridging chemotherapy in the CAR T-cell setting is not fully understood and subject to multiple confounding factors. To reduce the potential confounding due the differences in PICOS (population, intervention, comparator, outcomes, and study design) among the studies identified through our systematic review, we conducted a feasibility assessment prior to running the analyses and excluded four studies with substantially different PICOS.

Furthermore, methodological limitation stems from the fact that the MAIC method assumes all key prognostic factors differentially distributed across studies are taken into account (42,65). However, we could not adjust for all important prognostic factors since eligible studies reported different patient characteristics to describe their study samples, which limited the number of common key baseline covariates reported by experimental CAR T-cell common with ZUMA-1. For example, two important key baseline covariates for R-R DLBCL that lacked in experimental CAR T-cell trials were International Prognostic Index (IPI) and Eastern Cooperative Oncology Group (ECOG) Performance Status. Even when a similar patient characteristic was reported across studies, it was often measured by different scales in different papers, which makes the adjustment by MAIC impossible, such as International Prognostic Indexes (IPI), age-adjusted IPI and Revised-IPI. More serious inconsistency was found across the trials were the differential definition of relapsed disease as a baseline characteristic between ZUMA-1 and eligible IPD trials in this study. ZUMA-1 defined refractoriness as patients who had stable disease (SD) as their best response to the last line of therapy or those who had relapsed within 12 months of a consolidative ASCT. In IPD trials, besides using the same definition as that of the ZUMA-1 trial, progression at any time after the last line of therapy is also included as a criterion for relapsed disease. Therefore, we categorized patients as either refractory or relapsed in the MAICs irrespective of the type of relapse. Regarding the MAIC of safety outcomes, IPD trials used different grading systems for CRS and NT from ZUMA-1, which used the Lee criteria (66).

Hence, only a few important and mutually reported variables were adjusted for in the analysis. Consequently, any hidden difference in other unmeasured patient characteristics across studies could invalidate the findings. For example, pooled populations for the dual targeting and third generation CAR T-cells comprised of ethnically diverse patients within each pool consisting of the clinical trials conducted in China and USA (Table 1). To the best of our knowledge, there is no data available yet whether ethnicity has an impact on the CAR T-cell treatment outcomes. Apart from simple inverse weighting, advanced statistical methods based on doubly robust estimation have been developed to adjust for between-trial heterogeneity in patient characteristics (65). These approaches require specifying one model for the weight and another model for the outcome of interest. The advantage is that only one of these two models needs to be correctly specified to obtain valid and less biased results. Despite being more robust statistical solutions beyond the simple inverse weighting approach (used in MAIC), these methods were not used in the current study because theywere not feasible in our study due to the limited sample size. This study aims to build a basis for further exploration but not to draw definitive conclusions due to the small sample sizes of the contributing trials. Although a larger case series are needed to confirm these results, our findings are biologically plausible and clinically meaningful while corroborating with existing pre-clinical and clinical literature.

## CONCLUSION

In conclusion, the MAIC suggests a dual targeting approach using tandem CD19.CD20.4-1BB may have enhanced efficacy and safety compared to Yescarta. The hazard ratios of PFS were quantitatively in favor of the third generation, the sequential administration of ASCT and CD19.CD28 CAR T-cells and of the CD19. 4-1BBζ manufactured and evaluated among Chinese patients, although none of them reached statistical significance. The safety-enhanced CAR T-cell constructs included in our analysis, such as Hu19. CD8.28Z and CD19. BBz.86 demonstrated a remarkable safety profile with no occurrence of severe adverse events, yet without improvement in PFS compared to that of Yescarta.

While our results have shown the potential efficacy and safety advantages of tandem dual targeting approaches over the Yescarta, multi-targeted CAR T-cells, in general, are not likely to overcome the other resistance mechanisms beyond target antigen loss, such as the resistance involved with IL-6/STAT3 pathways, disruption of gene regulations for T-cell differentiation and exhaustion (67), and PD-L1 induced inhibition of CAR-T cells (68). Currently, a feasibility of multi-target CAR T-cells to mature into a routine clinical practice appear to be low given its higher production cost than single antigen targeting CARs owing to its multiple viral transductions and more complex manufacturing procedures. This study serves as a basis for further exploration and future studies shall aim to update the current study for longer follow-up data to identify the comparative efficacy, safety and feasibility of novel CAR T-cell products and the currently approved CAR T-cell therapies including Yescarta, Kymriah and most recently approved Lisocabtagene maraleucel (Breyanzi).

## Supporting information

Supplemental Figures, PRISMA flowchart and survival plots

## Data Availability

Individual patient data obtained from the eligible clinical trials analyzed in this study and any materials generated during the study are available and will be released via a material transfer agreement and study protocol, if available.

https://journal.ppcr.org/index.php/ppcrjournal/article/view/142

## ACKNOWLEDGMENTS

We would like to thank Professor Felipe Fregni, MD, Ph.D., MPH, Med and Alma Sanchez M.D (Harvard T.H. Chan School of Public Health), Paola A. Correa Ph.D (Howard Hughes Medical institute), and Tat-Thang Vo, Pharm.D, Ph.D (The Wharton School, University of Pennsylvania) for their expert advice.

## DATA AVAILABILITY

Ethical and informed consent statement

This study was based on the published data by the eligible clinical trials in this study. Therefore, ethics committee approval or informed consent statement were not required since the study did not involve patients or the public in the design, conduct, reporting, or dissemination plans.

## ABBREVIATIONS

ASCT: autologous stem cell transplantation
B-ALL: B-cell acute lymphoblastic leukemia
BCMA: B-cell maturation antigen
CAR: chimeric antigen receptor
CD: cluster of differentiation
ChiCTR: Chinese clinical trial registry
CI: confidence interval
CR: complete response
CRES: car T cell-related encephalopathy syndrome
CRR: complete response rate
CRS: cytokine release syndrome
CRS: cytokine release syndrome
DLBCL: diffuse large B-cell lymphoma
DOR: duration of response
EFS: event-free survival
ESS: effective sample size
HGBCL: high-grade B-cell lymphoma
HR: hazard ratio
Hu: human
IFN-γ: interferon-γ
IL-2: interleukins-2
IPD: individual patient data
KM: Kaplan-Meier
LBCL: large B-cell lymphoma
MAIC: matching adjusted indirect comparisons
NCT: national clinical trial
NHL: non-Hodgkin lymphoma
NT: neurotoxicity
OR: odds ratio
ORR: objective response rate
OS: overall survival
PFS: progression free survival
PH: proportional hazards
PPCR: principles and practice of clinical research
PRISMA: preferred reporting items for systematic reviews and meta-analyses
R/R LBCL: relapsed/refractory large B-cell lymphoma
RR: relative risk
RS: Richter’s transformation
scFv: single-chain variable fragment
SD: stable disease
TNF-α: tumor necrosis factor-α
trDLBCL: transformed DLBCL
ZUMA-1: name of Yescarta clinical trial

## REFERENCES

1. Morton LM, Wang SS, Devesa SS, Hartge P, Weisenburger DD, Linet MS. Lymphoma incidence patterns by WHO subtype in the United States, 1992-2001. Blood [Internet]. 2006;107(1):265–76. Available from: /pmc/articles/PMC1895348/

2. Howlader N, Noon AM KM. Cancer Statistics Review, 1975-2013 - Previous Version - SEER Cancer Statistics Review [Internet]. 2015 [cited 2021 Jun 9]. Available from: https://seer.cancer.gov/archive/csr/1975_2013/

3. Friedberg JW. Relapsed/refractory diffuse large B-cell lymphoma. [Internet]. Vol. 2011, Hematology / the Education Program of the American Society of Hematology. American Society of Hematology. Education Program. Hematology Am Soc Hematol Educ Program; 2011. p. 498–505. Available from: https://pubmed.ncbi.nlm.nih.gov/22160081/

4. Neste E Van Den, Schmitz N, Mounier N, Gill D, Linch D, Trneny M, et al. Outcome of patients with relapsed diffuse large B-cell lymphoma who fail second-line salvage regimens in the International CORAL study. Bone Marrow Transplant [Internet]. 2016;51(1):51–7. Available from: https://pubmed.ncbi.nlm.nih.gov/26367239/

5. Gisselbrecht C, Schmitz N, Mounier N, Gill DS, Linch DC, Trneny M, et al. Rituximab maintenance therapy after autologous stem-cell transplantation in patients with relapsed CD20+ diffuse large B-cell lymphoma: Final analysis of the collaborative trial in relapsed aggressive lymphoma. J Clin Oncol [Internet]. 2012;30(36):4462–9. Available from: https://pubmed.ncbi.nlm.nih.gov/23091101/

6. Abramson JS, Palomba ML, Gordon LI, Lunning MA, Wang M, Arnason J, et al. Lisocabtagene maraleucel for patients with relapsed or refractory large B-cell lymphomas (TRANSCEND NHL 001): a multicentre seamless design study. Lancet. 2020 Sep 19;396(10254):839–52.

7. Neelapu SS, Locke FL, Bartlett NL, Lekakis LJ, Miklos DB, Jacobson CA, et al. Axicabtagene Ciloleucel CAR T-Cell Therapy in Refractory Large B-Cell Lymphoma. N Engl J Med [Internet]. 2017 Dec 28 [cited 2021 May 13];377(26):2531–44. Available from: https://www.nejm.org/doi/full/10.1056/NEJMoa1707447

8. Schuster SJ, Bishop MR, Tam CS, Waller EK, Borchmann P, McGuirk JP, et al. Tisagenlecleucel in Adult Relapsed or Refractory Diffuse Large B-Cell Lymphoma. N Engl J Med [Internet]. 2019 Jan 3 [cited 2021 Jun 9];380(1):45–56. Available from: https://www.nejm.org/doi/full/10.1056/nejmoa1804980

9. Locke FL, Ghobadi A, Jacobson CA, Miklos DB, Lekakis LJ, Oluwole OO, et al. Long-term safety and activity of axicabtagene ciloleucel in refractory large B-cell lymphoma (ZUMA-1): a single-arm, multicentre, phase 1–2 trial. Lancet Oncol. 2019 Jan 1;20(1):31–42.

10. Ruella M, Barrett DM, Kenderian SS, Shestova O, Hofmann TJ, Perazzelli J, et al. Dual CD19 and CD123 targeting prevents antigen-loss relapses after CD19-directed immunotherapies. J Clin Invest. 2016 Oct 3;126(10):3814–26.

11. Hamieh M, Dobrin A, Cabriolu A, van der Stegen SJC, Giavridis T, Mansilla-Soto J, et al. CAR T cell trogocytosis and cooperative killing regulate tumour antigen escape. Nature [Internet]. 2019 Apr 4 [cited 2021 Jun 2];568(7750):112–6. Available from: https://pubmed.ncbi.nlm.nih.gov/30918399/

12. Shah NN, Maatman T, Hari P, Johnson B. Multi targeted CAR-T cell therapies for B-cell malignancies. Front Oncol. 2019;9(MAR):1–7.

13. Enblad G, Karlsson H, Gammelgård G, Wenthe J, Lövgren T, Amini RM, et al. A phase I/IIa trial using CD19-targeted third-generation CAR T cells for lymphoma and leukemia. Clin Cancer Res. 2018;24(24):6185–94.

14. Huang C, Wu L, Liu R, Li W, Li Z, Li J, et al. Efficacy and safety of CD19 chimeric antigen receptor T cells in the treatment of 11 patients with relapsed/refractory B-cell lymphoma: a single-center study. Ann Transl Med Vol 8, No 17 (September 2020) Ann Transl Med [Internet]. 2020; Available from: http://atm.amegroups.com/article/view/49940

15. Ramos CA, Rouce R, Robertson CS, Reyna A, Narala N, Vyas G, et al. In Vivo Fate and Activity of Second-versus Third-Generation CD19-Specific CAR-T Cells in B Cell Non-Hodgkin’s Lymphomas. Mol Ther. 2018 Dec 5;26(12):2727–37.

16. Brudno JN, Lam N, Vanasse D, Shen Y wei, Rose JJ, Rossi J, et al. Safety and feasibility of anti-CD19 CAR T cells with fully human binding domains in patients with B-cell lymphoma. Nat Med [Internet]. 2020 Feb 1 [cited 2020 Nov 10];26(2):270–80. Available from: https://doi.org/10.1038/s41591-019-0737-3

17. Ying Z, Huang XF, Xiang X, Liu Y, Kang X, Song Y, et al. A safe and potent anti-CD19 CAR T cell therapy. Nat Med [Internet]. 2019 Jun 1 [cited 2020 Nov 18];25(6):947–53. Available from: https://www-nature-com.libproxy.albany.edu/articles/s41591-019-0421-7

18. Cao Y, Lu W, Sun R, Jin X, Cheng L, He X, et al. Anti-CD19 Chimeric Antigen Receptor T Cells in Combination With Nivolumab Are Safe and Effective Against Relapsed/Refractory B-Cell Non-hodgkin Lymphoma. Front Oncol [Internet]. 2019 Aug 19 [cited 2020 Nov 25];9(AUG):767. Available from: https://www.frontiersin.org/article/10.3389/fonc.2019.00767/full

19. Kebriaei P, Singh H, Huls MH, Figliola MJ, Bassett R, Olivares S, et al. Phase i trials using sleeping beauty to generate CD19-specific CAR T cells. J Clin Invest [Internet]. 2016 Sep 1 [cited 2020 Oct 15];126(9):3363–76. Available from: http://www.imgt.org/

20. Sauter CS, Senechal B, Rivière I, Ni A, Bernal Y, Wang X, et al. CD19 CAR T cells following autologous transplantation in poor-risk relapsed and refractory B-cell non-Hodgkin lymphoma. Blood [Internet]. 2019 Aug [cited 2020 Oct 15];134(7):626–35. Available from: https://pubmed.ncbi.nlm.nih.gov/31262783/

21. Wang X, Popplewell LL, Wagner JR, Naranjo A, Blanchard MS, Mott MR, et al. Phase 1 studies of central memory-derived CD19 CAR T-cell therapy following autologous HSCT in patients with B-cell NHL. Blood [Internet]. 2016 Jun 16 [cited 2020 Oct 15];127(24):2980–90. Available from: http://ashpublications.org/blood/article-pdf/127/24/2980/1394503/2980.pdf

22. Schneider D, Xiong Y, Wu D, Hu P, Alabanza L, Steimle B, et al. Trispecific CD19-CD20-CD22-targeting duoCAR-T cells eliminate antigen-heterogeneous B cell tumors in preclinical models. Sci Transl Med [Internet]. 2021 Mar 24 [cited 2021 May 23];13(586). Available from: https://stm.sciencemag.org/content/13/586/eabc6401

23. Schneider D, Xiong Y, Wu D, Nölle V, Schmitz S, Haso W, et al. A tandem CD19/CD20 CAR lentiviral vector drives on-target and off-target antigen modulation in leukemia cell lines. J Immunother Cancer. 2017;5(1):1–17.

24. Zah E, Lin MY, Anne SB, Jensen MC, Chen YY. T cells expressing CD19/CD20 bispecific chimeric antigen receptors prevent antigen escape by malignant B cells. Cancer Immunol Res [Internet]. 2016 Jun 1 [cited 2021 May 13];4(6):498–508. Available from: http://www.aacrjournals.org

25. Ormhøj M, Scarfo I, Cabral ML, Bailey SR, Lorrey SJ, Bouffard AA, et al. Chimeric antigen receptor T cells targetin CD79b show efficacy in lymphoma with or without cotargeting CD19. Clin Cancer Res [Internet]. 2019 Dec 1 [cited 2021 May 13];25(23):7046–57. Available from: http://clincancerres.aacrjournals.org/

26. Mihara K, Yanagihara K, Takigahira M, Kitanaka A, Imai C, Bhattacharyya J, et al. Synergistic and persistent effect of T-cell immunotherapy with anti-CD19 or anti-CD38 chimeric receptor in conjunction with rituximab on B-cell non-Hodgkin lymphoma. Br J Haematol [Internet]. 2010 Oct 1 [cited 2021 May 13];151(1):37–46. Available from: http://symatlas.gnf.org

27. Scarfò I, Ormhøj M, Frigault MJ, Castano AP, Lorrey S, Bouffard AA, et al. Anti-CD37 chimeric antigen receptor T cells are active against B-A nd T-cell lymphomas. Blood [Internet]. 2018 Oct 4 [cited 2021 May 13];132(14):1495–506. Available from: http://ashpublications.org/blood/article-pdf/132/14/1495/1727274/blood842708.pdf

28. Zhang Y. Safety and efficacy of optimized tandem CD19/CD20 CAR-engineered T cells in patients with relapsed/refractory non-Hodgkin lymphoma. J Clin Oncol. 2020 May 20;38(15_suppl):3034–3034.

29. Tong C, Zhang Y, Liu Y, Ji X, Zhang W-Y, Guo Y, et al. Optimized tandem CD19/CD20 CAR-engineered T cells in refractory/relapsed B cell lymphoma. Blood [Internet]. 2020;((Tong C.; Zhang Y.; Liu Y.; Han X.; Ti D.; Wang C.; Wu Z.; Han W.) Chinese PLA General Hospital, China). Available from: https://www.embase.com/search/results?subaction=viewrecord&id=L632129081&from=export

30. Shah NN, Johnson BD, Schneider D, Zhu F, Szabo A, Keever-Taylor CA, et al. Bispecific anti-CD20, anti-CD19 CAR T cells for relapsed B cell malignancies: a phase 1 dose escalation and expansion trial. Nat Med. 2020;26(10):1569–75.

31. Sang W, Shi M, Yang J, Cao J, Xu L, Yan D, et al. Phase II trial of co-administration of CD19-and CD20-targeted chimeric antigen receptor T cells for relapsed and refractory diffuse large B cell lymphoma. Cancer Med. 2020 Jul;9(16):5827–38.

32. Zeng C, Cheng J, Li T, Huang J, Li C, Jiang L, et al. Efficacy and toxicity for CD22/CD19 chimeric antigen receptor T-cell therapy in patients with relapsed/refractory aggressive B-cell lymphoma involving the gastrointestinal tract. Cytotherapy. 2020 Mar 1;22(3):166–71.

33. Wang N, Hu X, Cao W, Li C, Xiao Y, Cao Y, et al. Efficacy and safety of CAR19/22 T-cell cocktail therapy in patients with refractory/relapsed B-cell malignancies. Blood [Internet]. 2020 Jan 2 [cited 2021 May 13];135(1):17–27. Available from: http://www.chictr.org.cn

34. Zhong XS, Matsushita M, Plotkin J, Riviere I, Sadelain M. Chimeric antigen receptors combining 4-1BB and CD28 signaling domains augment PI 3 kinase/AKT/Bcl-X L activation and CD8 T cell-mediated tumor eradication. Mol Ther. 2010 Feb 1;18(2):413–20.

35. Li C, Zhang Y, Zhang C, Chen J, Lou X, Chen X, et al. Comparison of CAR-T19 and autologous stem cell transplantation for refractory/relapsed non-Hodgkin’s lymphoma. JCI Insight [Internet]. 2019 Aug 22 [cited 2021 May 20];4(17). Available from: /pmc/articles/PMC6777912/

36. Wang Y, Zhang W ying, Han Q wang, Liu Y, Dai H ren, Guo Y lei, et al. Effective response and delayed toxicities of refractory advanced diffuse large B-cell lymphoma treated by CD20-directed chimeric antigen receptor-modified T cells. Clin Immunol. 2014 Dec 1;155(2):160–75.

37. Zhang WY, Liu Y, Wang Y, Wang CM, Yang QM, Zhu HL, et al. Long-term safety and efficacy of CART-20 cells in patients with refractory or relapsed b-cell non-hodgkin lymphoma: 5-years follow-up results of the phase i and iia trials. Signal Transduct Target Ther. 2017;2:0–3.

38. Zhang WY, Wang Y, Guo YL, Dai HR, Yang QM, Zhang YJ, et al. Treatment of cd20-directed chimeric antigen receptor-modified t cells in patients with relapsed or refractory b-cell non-hodgkin lymphoma: An early phase iia trial report. Signal Transduct Target Ther. 2016;1(January):1–9.

39. Borchmann P. Phase I Trial of MB-CART2019.1, a Novel CD20 and CD19 Targeting Tandem Chimeric Antigen Receptor, in Patients with Relapsed or Refractory B-Cell Non-Hodgkin Lymphoma. ASH; 2020.

40. Shadman M, Gopal AK, Smith SD, Lynch RC, Ujjani CS, Turtle CJ, et al. CD20 Targeted CAR-T for High-Risk B-Cell Non-Hodgkin Lymphomas. Blood. 2019 Nov 13;134(Supplement_1):3235–3235.

41. Salles G, Barrett M, Foà R, Maurer J, O’Brien S, Valente N, et al. Rituximab in B-Cell Hematologic Malignancies: A Review of 20 Years of Clinical Experience [Internet]. Vol. 34, Advances in Therapy. Springer Healthcare; 2017 [cited 2021 May 22]. p. 2232–73. Available from: http://www.medengine.com/Redeem/

42. Phillippo DM, Ades AE, Dias S, Palmer S, Abrams KR, Welton NJ. Methods for Population-Adjusted Indirect Comparisons in Health Technology Appraisal. Med Decis Mak. 2018 Feb 1;38(2):200–11.

43. Signorovitch JE, Sikirica V, Erder MH, Xie J, Lu M, Hodgkins PS, et al. Matching-adjusted indirect comparisons: A new tool for timely comparative effectiveness research. Value Heal. 2012 Sep 1;15(6):940–7.

44. NIH. Study Quality Assessment Tools | NHLBI, NIH [Internet]. NIH. 2013 [cited 2021 Oct 24]. Available from: https://www.nhlbi.nih.gov/health-topics/study-quality-assessment-tools

45. Weinstein B, Muresan B, Solano S, Vaz de Macedo A, Lee Y. Efficacy and Safety of Experimental versus Approved CAR T-cell Therapies in Large B-cell Lymphoma Using Matching Adjusted Indirect Comparisons: A. http://journal.ppcr.org [Internet]. [cited 2021 Jun 9]; Available from: https://journal.ppcr.org/index.php/ppcrjournal/article/view/142

46. Guyot P, Ades AE, Ouwens MJNM, Welton NJ. Enhanced secondary analysis of survival data: Reconstructing the data from published Kaplan-Meier survival curves. BMC Med Res Methodol [Internet]. 2012 Feb 1 [cited 2021 Jun 9];12(1):1–13. Available from: https://link.springer.com/articles/10.1186/1471-2288-12-9

47. Oluwole OO, Jansen JP, Lin VW, Chan K, Keeping S, Navale L, et al. Comparing Efficacy, Safety, and Preinfusion Period of Axicabtagene Ciloleucel versus Tisagenlecleucel in Relapsed/Refractory Large B Cell Lymphoma: Comparative Study of Axicabtagene Ciloleucel and Tisagenlecleucel. Biol Blood Marrow Transplant [Internet]. 2020 Sep 1 [cited 2021 May 2];26(9):1581–8. Available from: https://pubmed.ncbi.nlm.nih.gov/32561336/

48. Chen X, Li X, Liu Y, Zhang Z, Zhang X, Huang J, et al. A Phase I clinical trial of chimeric antigen receptor-modified T cells in patients with relapsed and refractory lymphoma. Immunotherapy. 2020 Jul 1;12(10):681–96.

49. Wang T, Gao L, Wang Y, Zhu W, Xu L, Wang Y, et al. Hematopoietic stem cell transplantation and chimeric antigen receptor T cell for relapsed or refractory diffuse large B-cell lymphoma. Immunotherapy [Internet]. 2020 Sep 1 [cited 2020 Oct 15];12(13):997–1006. Available from: https://www.futuremedicine.com/doi/abs/10.2217/imt-2020-0075

50. Guo Z, Tu S, Yu S, Wu L, Pan W, Chang N, et al. Preclinical and clinical advances in dual-target chimeric antigen receptor therapy for hematological malignancies [Internet]. Vol. 112, Cancer Science. Blackwell Publishing Ltd; 2021 [cited 2021 May 14]. p. 1357–68. Available from:/pmc/articles/PMC8019219/

51. Shah NN, Fry TJ. Mechanisms of resistance to CAR T cell therapy. Nat Rev Clin Oncol [Internet]. 2019;16(6):372–85. Available from: http://dx.doi.org/10.1038/s41571-019-0184-6

52. Hossain N, Sahaf B, Abramian M, Spiegel JY, Kong K, Kim S, et al. Phase I Experience with a Bi-Specific CAR Targeting CD19 and CD22 in Adults with B-Cell Malignancies. Blood. 2018 Nov 29;132(Supplement 1):490–490.

53. Amrolia PJ, Wynn R, Hough RE, Vora A, Bonney D, Veys P, et al. Phase I Study of AUTO3, a Bicistronic Chimeric Antigen Receptor (CAR) T-Cell Therapy Targeting CD19 and CD22, in Pediatric Patients with Relapsed/Refractory B-Cell Acute Lymphoblastic Leukemia (r/r B-ALL): Amelia Study. Blood. 2019 Nov 13;134(Supplement_1):2620–2620.

54. Yang J, Li J, Zhang X, Lv F, Guo X, Wang Q, et al. A Feasibility and Safety Study of CD19 and CD22 Chimeric Antigen Receptors-Modified T Cell Cocktail for Therapy of B Cell Acute Lymphoblastic Leukemia. Blood. 2018 Nov 29;132(Supplement 1):277–277.

55. Carpenito C, Milone MC, Hassan R, Simonet JC, Lakhal M, Suhoski MM, et al. Control of large, established tumor xenografts with genetically retargeted human T cells containing CD28 and CD137 domains. Proc Natl Acad Sci U S A [Internet]. 2009 Mar 3 [cited 2021 May 16];106(9):3360–5. Available from: https://pubmed.ncbi.nlm.nih.gov/19211796/

56. Gomes-Silva D, Mukherjee M, Srinivasan M, Krenciute G, Dakhova O, Zheng Y, et al. Tonic 4-1BB Costimulation in Chimeric Antigen Receptors Impedes T Cell Survival and Is Vector-Dependent. Cell Rep. 2017 Oct;21(1):17–26.

57. Feucht J, Sun J, Eyquem J, Ho YJ, Zhao Z, Leibold J, et al. Calibration of CAR activation potential directs alternative T cell fates and therapeutic potency. Nat Med [Internet]. 2019 Jan 1 [cited 2021 May 20];25(1):82–8. Available from: https://pubmed.ncbi.nlm.nih.gov/30559421/

58. Majzner RG, Rietberg SP, Sotillo E, Dong R, Vachharajani VT, Labanieh L, et al. Tuning the antigen density requirement for car T-cell activity. Cancer Discov [Internet]. 2020 May 1 [cited 2021 May 20];10(5):702–23. Available from: http://cancerdiscovery.aacrjournals.org/

59. Philip T, Guglielmi C, Hagenbeek A, Somers R, Van Der Lelie H, Bron D, et al. Autologous Bone Marrow Transplantation as Compared with Salvage Chemotherapy in Relapses of Chemotherapy-Sensitive Non-Hodgkin’s Lymphoma. N Engl J Med [Internet]. 1995 Dec 7 [cited 2021 May 21];333(23):1540–5. Available from: https://pubmed.ncbi.nlm.nih.gov/7477169/

60. June CH, Sadelain M. Chimeric Antigen Receptor Therapy. N Engl J Med [Internet]. 2018 Jul 5 [cited 2021 May 23];379(1):64–73. Available from: http://www.nejm.org/doi/10.1056/NEJMra1706169

61. Horna P, Nowakowski G, Endell J, Boxhammer R. Comparative Assessment of Surface CD19 and CD20 Expression on B-Cell Lymphomas from Clinical Biopsies: Implications for Targeted Therapies. Blood. 2019 Nov 13;134(Supplement_1):5345–5345.

62. Majzner RG, Mackall CL. Clinical lessons learned from the first leg of the CAR T cell journey [Internet]. Vol. 25, Nature Medicine. Nature Publishing Group; 2019 [cited 2021 May 17]. p. 1341–55. Available from: https://www-nature-com.libproxy.albany.edu/articles/s41591-019-0564-6

63. Maloney DG, Kuruvilla J, Fox CP, Cartron G, Li D, Hasskarl J, et al. Matching-Adjusted Indirect Comparison (MAIC) of Lisocabtagene Maraleucel (liso-cel) Vs Axicabtagene Ciloleucel (axi-cel) and Tisagenlecleucel in Relapsed/Refractory (R/R) Large B-Cell Lymphoma (LBCL). Blood. 2020 Nov 5;136(Supplement 1):18–9.

64. Ramsborg CG, Guptill P, Weber C, Christin B, Larson RP, Lewis K, et al. JCAR017 Is a Defined Composition CAR T Cell Product with Product and Process Controls That Deliver Precise Doses of CD4 and CD8 CAR T Cell to Patients with NHL. Blood. 2017 Dec 7;130(Supplement 1):4471–4471.

65. Vo TT, Porcher R, Chaimani A, Vansteelandt S. A novel approach for identifying and addressing case-mix heterogeneity in individual participant data meta-analysis. Res Synth Methods [Internet]. 2019 Dec 1 [cited 2021 Jun 14];10(4):582–96. Available from: https://pubmed.ncbi.nlm.nih.gov/31682071/

66. Lee DW, Santomasso BD, Locke FL, Ghobadi A, Turtle CJ, Brudno JN, et al. ASTCT Consensus Grading for Cytokine Release Syndrome and Neurologic Toxicity Associated with Immune Effector Cells. Vol. 25, Biology of Blood and Marrow Transplantation. Elsevier Inc.; 2019. p. 625–38.

67. Fraietta JA, Lacey SF, Orlando EJ, Pruteanu-Malinici I, Gohil M, Lundh S, et al. Determinants of response and resistance to CD19 chimeric antigen receptor (CAR) T cell therapy of chronic lymphocytic leukemia. Nat Med 2018 245 [Internet]. 2018 Apr 30 [cited 2021 Oct 24];24(5):563–71. Available from: https://www.nature.com/articles/s41591-018-0010-1

68. Rafiq S, Yeku OO, Jackson HJ, Purdon TJ, van Leeuwen DG, Drakes DJ, et al. Targeted delivery of a PD-1-blocking scFv by CAR-T cells enhances anti-tumor efficacy in vivo. Nat Biotechnol 2018 369 [Internet]. 2018 Oct 1 [cited 2021 Oct 24];36(9):847–56. Available from: https://www.nature.com/articles/nbt.4195

