## Supplemental Figures, PRISMA flowchart and survival plots for "Matching Adjusted Indirect Comparisons (MAICs) and Systematic Review: Efficacy and Safety of Experimental Chimeric Antigen Receptor (CAR) T-cells versus Axicabtagene ciloleucel (Yescarta) for the Treatment of Relapsed/Refractory Large B-Cell Lymphoma (LBCL)"

**Figure S1.** PRISMA Flow Diagram, Large B-Cell Lymphoma (LBCL)

### Eligibility

### Screening

### Identification

**N=12** records from ChiCTR

and clinicaltrials.gov

**N=308** records identified through e-database searching

**N=15** studies used for quantitative synthesis

**N=106** excluded

49 Non-LBCL studies

15 Observational studies

14 Meta-analyses

12 Abstracts

11 Conference papers

5 Opinion letters

**N=161** records after duplicates removed

**N=25** excluded

3 Pivotal trials

12 Case reports

5 Case-series

3 Letter to editor

2 No IPD data

**N=55** full-text articles screened

**N=30** full-text articles assessed for eligibility

**N=9** excluded

3 Sample size <10

2 Only 28-day follow-up

4 studies with ineligible PICOS*

**N=24** studies used in feasibility assessment

### Included

*PICOS - population, intervention, comparator, outcomes, and study design

**Figure S2.** MAIC of experimental CAR T-cells and Yescarta regarding PFS among patients who received infusion. Kaplan Meier survival curves. Hazard Ratios and 95% Confidence Intervals computed through Cox Proportional Hazards Models.

|  | Months |
| --- | --- |
| A. Dual targeting  Tandem CD19. CD20 with 4-1BBζ | 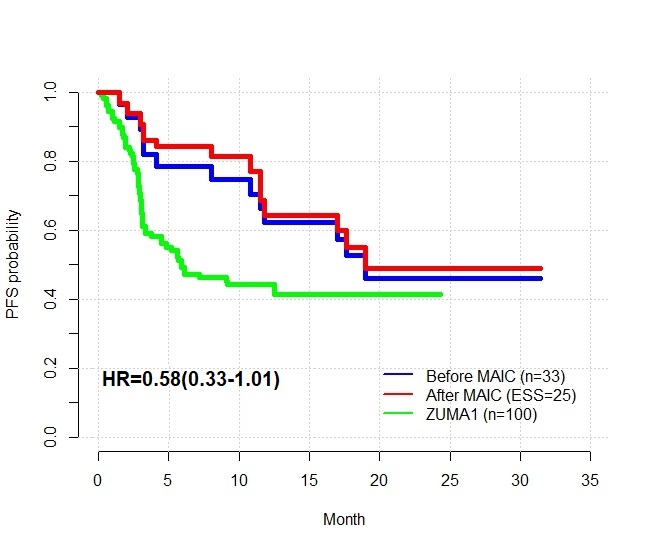 |
| Co-infusion CD19 & CD20 with 4-1BBζ | 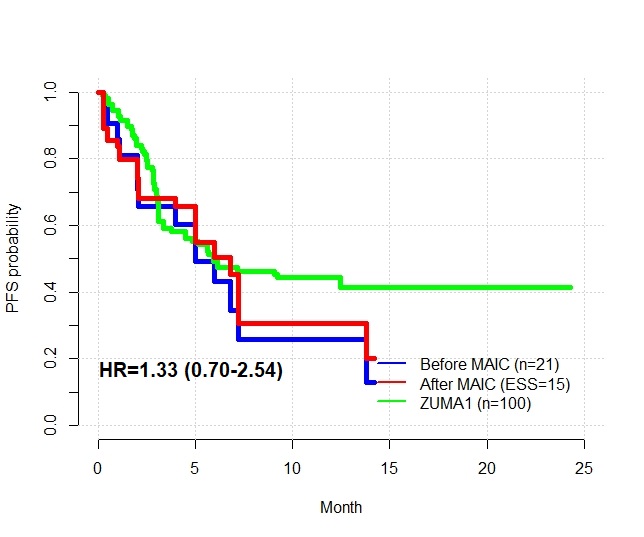 |
| B. Third generation CARs  CD19 with CD28ζ & 4-1BBζ | 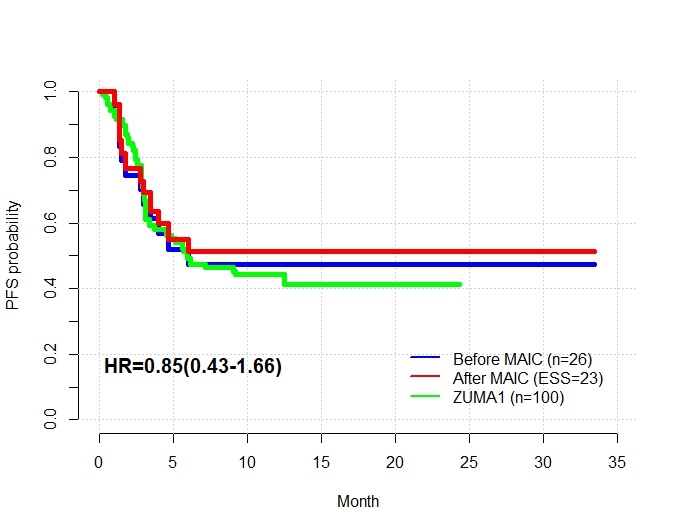 |
| C. Sequential administration of ASCT and CD19.CD28ζ | 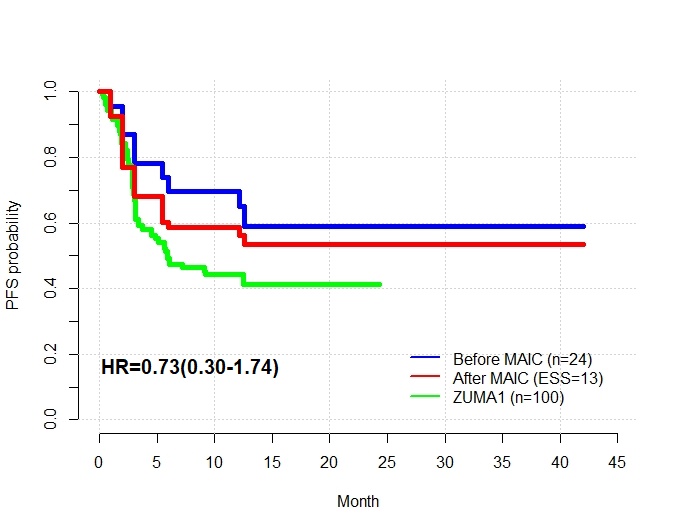 |
| D. Modified co-stimulatory domain for reduced toxicity:  Hu19.CD8.28Z | 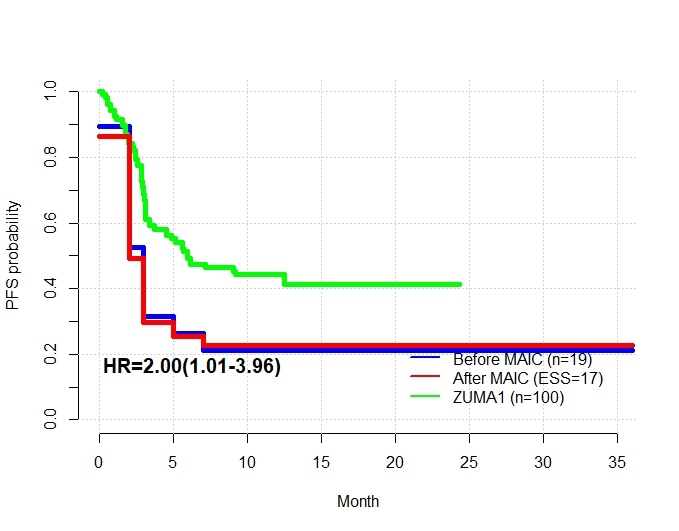 |
| CD19. BBz.86 | 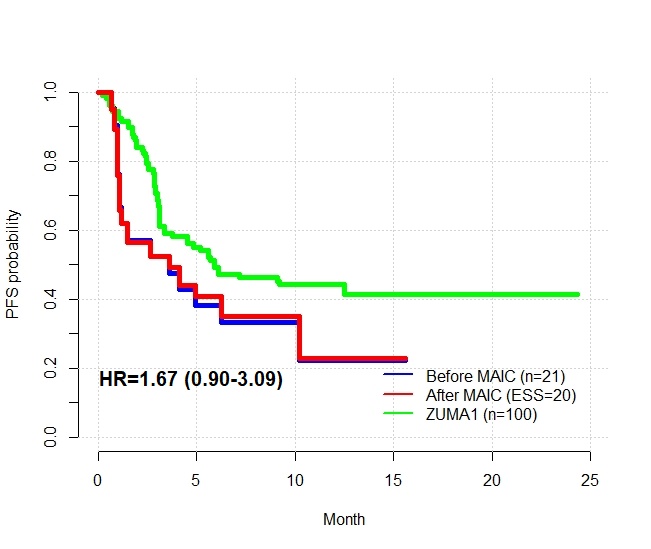 |
| E. Alternative target antigen  CD20. 4-1BBζ | 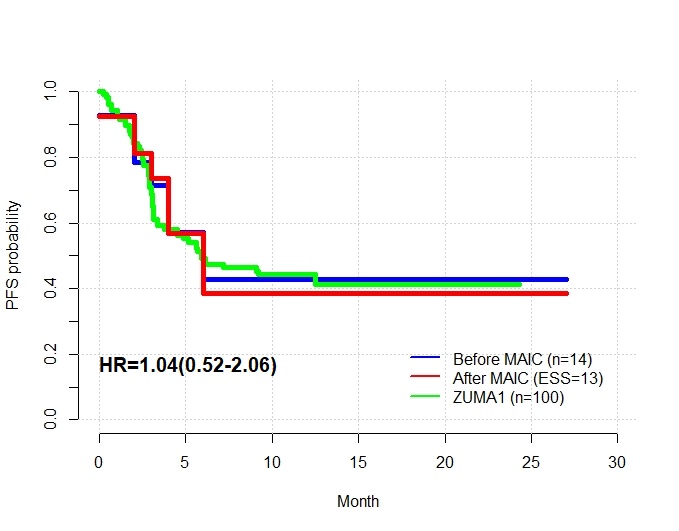 |
| F. Alternative co-stimulatory domain  CD19. 4-1BBζ | 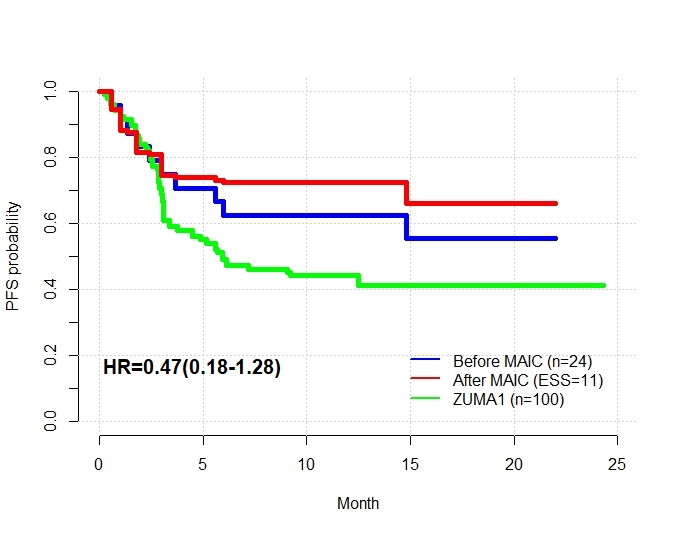 |
